## Supplementary Material for "High-dose vitamin D_3_ in the treatment of complicated severe acute malnutrition in Pakistan: a double-blind randomised controlled trial (ViDiSAM)"

### **Supplementary Methods**

#### Randomisation procedure

Eligible participants were individually randomised to intervention vs. control arms with a one-to-one allocation ratio and stratification by hospital of recruitment as follows: prior to the start of recruitment, the trial statistician (NF) prepared one hospital randomisation list and two separate participant randomisation lists (one for each site). The hospital randomisation list comprised one pair of letters per participating hospital (Ganga Ram Hospital: A/B, THQ Hospital: Y/Z). One letter within each pair was randomly assigned to the active arm of the trial, and the other was randomly assigned to the placebo arm, using a computer-generated random sequence. The resulting hospital randomisation list was sent to GT Pharma Ltd, Lahore, Pakistan (the manufacturer of the investigational medicinal product [IMP]), with a copy retained by the trial statistician. Neither participants nor trial staff had access to this list. GT Pharma then used this hospital randomisation list to label a total of 600 glass ampoules (300 each containing 5 mg vitamin D_3_ in 1 ml ethyl oleate, and 300 each containing 1 ml ethyl oleate without vitamin D_3_). The label on each ampoule clearly displayed one of the letters A/B/Y/Z (the randomisation codes indicating whether the ampoule contained active or placebo IMP). Labelled ampoules were then packed into boxes according to the letter code displayed on their label, with each box labelled with the letter corresponding to the contents of the ampoules it contained. Two participant randomisation lists were then generated – one for each hospital site. Each of the participant randomisation lists comprised 999 4-digit numbers consisting of a 1-digit hospital identifier (1 for the Ganga Ram Hospital list, 3 for THQ Hospital), followed by a 3-digit participant identifier from 001 to 999 (e.g. 1-001, 1-002…; 3-001, 3-002 …). These 4-digit numeric codes were then randomly assigned to one or other of the one-letter randomisation codes in use for the corresponding hospital using a computer-generated random sequence employing a blocking structure. When eligibility of a new participant was confirmed (i.e. when the baseline serum albumin-adjusted calcium result was found to be ≤2.65 mmol/L), the participant randomisation list for the relevant site was used to assign IMP to each participant, based on the numeric component of their ID number.

### **Table S1.** Reasons for ineligibility to participate (n=306 participants screened)

| **Reason for ineligibility** | **No. of children^(1)^** |
| --- | --- |
| Consent not given | 28 |
| Aged < 6 or >59 months | 5 |
| Family moved / planned to move out of the study area within 6 months of enrolment | 29 |
| SAM without complications or MAM (moderate acute malnutrition) | 187 |
| Ingested a dose of vitamin D*>*200,000 IU (5 mg) in the 3 months prior to enrolment | 17 |
| Known diagnosis of primary hyperparathyroidism or sarcoidosis (i.e., conditions pre-disposing to vitamin D hypersensitivity) | 0 |
| Known neurodevelopmental disorder (e.g., cerebral palsy) | 23 |
| HIV infection | 5 |
| Taking anti-tuberculosis treatment | 7 |
| Unable to assess child’s developmental status at baseline using the Malawi Developmental Assessment Tool | 0 |
| Clinical signs of rickets | 12 |
| Baseline corrected serum calcium concentration >2.65 mmol/L | 7 |

1, total in this column adds to n=320 because 7 children fulfilled two ineligibility criteria simultaneously

### **Table S2.** Doses of study preparation received, by allocation (n=259)

| **Total number of doses received** | **Proportion receiving no. of doses, placebo arm** | **Proportion receiving no. of doses, vitamin D arm** |
| --- | --- | --- |
| **0** | 1/131 (0.8%) | 0/128 (0.0%) |
| **1** | 2/131 (1.5%) | 1/128 (0.8%) |
| **2** | 128/131 (97.7%) | 127/128 (99.2%) |

### **Table S3.** Sub-group analysis: major efficacy outcomes at 2-month follow-up by baseline weight-for-height or -length z-score

| **Outcome** | **Baseline weight-for-height or -length z-score** | **Placebo, mean value (s.d.) [n]** | **Vitamin D, mean value (s.d.) [n]** | **Adjusted mean difference (95% CI) *** | **P value** | **P for interaction (95% CI)** |
| --- | --- | --- | --- | --- | --- | --- |
| **Anthropometric/Clinical Outcomes** | | | | | | |
| Weight-for-height or -length z-score | ≥-3.0 | -1.78 (0.96), [31] | -1.86 (0.81), [31] | -0.12 (-0.45, 0.21) | 0.48 | 0.015 |
|  | <-3.0 | -2.84 (1.01), [95] | -2.83 (1.08), [89] | 0.05 (-0.19, 0.29) | 0.67 |  |
| Lean mass index | ≥-3.0 | 13.03 (2.16), [31] | 13.05 (2.24), [31] | 0.65 (-0.20, 1.51) | 0.14 | 0.47 |
|  | <-3.0 | 12.50 (2.10), [95] | 12.54 (2.41), [89] | -0.04 (-0.62, 0.54) | 0.90 |  |
| Mean mid-upper arm circumference, cm | ≥-3.0 | 11.69 (0.85), [31] | 11.88 (0.78), [31] | 0.04 (-0.28, 0.35) | 0.82 | 0.68 |
|  | <-3.0 | 11.51 (0.91), [95] | 11.54 (0.92), [89] | 0.07 (-0.16, 0.31) | 0.53 |  |
| Weight-for-age z-score | ≥-3.0 | -2.99 (1.23), [31] | -2.96 (0.99), [31] | -0.22 (-0.52, 0.08) | 0.14 | 0.14 |
|  | <-3.0 | -3.67 (1.03), [95] | -3.65 (1.12), [89] | 0.08 (-0.13, 0.28) | 0.45 |  |
| **Neurodevelopmental Outcomes** | | | | | | |
| Overall MDAT score | ≥-3.0 | 60.48 (21.56), [31] | 60.71 (12.78), [31] | -1.05 (-3.20, 1.10) | 0.34 | 0.43 |
|  | <-3.0 | 62.03 (21.54), [95] | 59.37 (23.21), [89] | -0.21 (-1.75, 1.34) | 0.79 |  |

*Adjusted for site of recruitment and baseline value. Abbreviations: MDAT: Malawi Developmental Assessment Tool, S.d.: Standard deviation, CI: Confidence interval.

### **Table S4.** Sub-group analysis: major efficacy outcomes at 2-month follow-up by sex

| **Outcome** | **Sex** | **Placebo, mean value (s.d.) [n]** | **Vitamin D, mean value (s.d.) [n]** | **Adjusted mean difference (95% CI) *** | **P value** | **P for interaction (95% CI)** |
| --- | --- | --- | --- | --- | --- | --- |
| **Anthropometric/Clinical Outcomes** | | | | | | |
| Weight-for-height/length z-score | Female | -2.43 (1.09), [72] | -2.34 (1.09), [71] | 0.03 (-0.23, 0.28) | 0.83 | 0.77 |
|  | Male | -2.79 (1.08), [56] | -2.90 (1.05), [52] | -0.06 (-0.40, 0.28) | 0.72 |  |
| Lean mass index | Female | 12.39 (2.08), [72] | 12.26 (2.02), [71] | -0.07 (-0.67, 0.52) | 0.81 | 0.22 |
|  | Male | 12.89 (2.13), [56] | 13.19 (2.67), [52] | 0.27 (-0.51, 1.05) | 0.50 |  |
| Mean mid-upper arm circumference, cm | Female | 11.50 (0.93), [72] | 11.65 (0.77), [71] | 0.13 (-0.10, 0.37) | 0.26 | 0.84 |
|  | Male | 11.67 (0.88), [56] | 11.63 (1.04), [52] | -0.02 (-0.33, 0.30) | 0.92 |  |
| Weight-for-age z-score | Female | -3.35 (1.12), [72] | -3.24 (1.09), [71] | 0.03 (-0.18, 0.24) | 0.81 | 0.93 |
|  | Male | -3.69 (1.08), [56] | -3.79 (1.08), [52] | -0.02 (-0.31, 0.26) | 0.87 |  |
| **Neurodevelopmental Outcomes** | | | | | | |
| Overall MDAT score | Female | 60.43 (19.66), [72] | 62.62 (20.36), [71] | 0.21 (-1.54, 1.96) | 0.82 | 0.22 |
|  | Male | 64.77 (25.39), [56] | 58.04 (23.74), [52] | -1.18 (-2.95, 0.58) | 0.19 |  |

*Adjusted for site of recruitment and baseline value. Abbreviations: MDAT: Malawi Developmental Assessment Tool, S.d.: Standard deviation, CI: Confidence interval.

### **Table S5.** Sub-group analysis: major efficacy outcomes at 2-month follow-up by baseline vitamin D status

| **Outcome** | **Baseline 25(OH)D concentration, nmol/L** | **Placebo, mean value (s.d.), [n]** | **Vitamin D, mean value (s.d.), [n]** | **Adjusted mean difference (95% CI)^[1]^** | **P for interaction (95% CI)** |
| --- | --- | --- | --- | --- | --- |
| **Anthropometric/Clinical Outcomes** | | | | | |
| Weight-for-height/length z-score | <50 | -2.68 (1.09), [36] | -2.50 (1.08), [31] | 0.11 (-0.32, 0.53) | 0.96 |
|  | ≥50 | -3.30 (1.14), [16] | -2.63 (1.05), [23] | 0.52 (0.02, 1.01) |  |
| Lean mass index, Ω^-1^ | <50 | 12.88 (2.02), [36] | 13.15 (2.87), [31] | 0.36 (-0.64, 1.37) | 0.23 |
|  | ≥50 | 13.26 (3.10), [16] | 12.90 (2.25), [23] | -0.42 (-1.73, 0.89) |  |
| Mean mid-upper arm circumference, cm | <50 | 11.64 (0.82), [36] | 11.70 (0.84), [31] | 0.08 (-0.26, 0.41) | 0.72 |
|  | ≥50 | 11.08 (1.34), [16] | 11.74 (0.98), [23] | 0.47 (-0.08, 1.02) |  |
| Weight-for-age z-score | <50 | -3.62 (1.06), [36] | -3.43 (1.10), [31] | 0.09 (-0.24, 0.43) | 0.44 |
|  | ≥50 | -3.90 (1.26), [16] | -3.61 (1.25), [23] | 0.16 (-0.25, 0.57) |  |
| **Neurodevelopmental Outcomes** | | | | | |
| Overall MDAT score | <50 | 59.14 (21.29), [36] | 59.55 (17.91), [31] | 0.45 (-2.10, 3.04) | 0.38 |
|  | ≥50 | 55.38 (20.55), [16] | 59.30 (25.19), [23] | 0.21 (-3.32, 3.75) |  |

1, Adjusted for site of recruitment and baseline value. Abbreviations: MDAT: Malawi Developmental Assessment Tool, S.d.: Standard deviation, CI: Confidence interval.

### **Table S6**. Serious adverse events by allocation: line listing

|  | **Placebo arm** | **Vitamin D arm** |
| --- | --- | --- |
| Hospitalisation for bronchopneumonia | 2 | 0 |
| Hospitalisation for croup | 1 | 0 |
| Hospitalisation for sepsis | 1 | 0 |
| Hospitalisation for gastroenteritis | 6^[1]^ | 3 |
| Hospitalisation for viral myocarditis | 1^[2]^ | 0 |
| **Total** | **11** | **3** |

1, of which one was fatal. 2, fatal event
